# Cost analysis of overseas versus domestic vaccination of US-bound refugees

**DOI:** 10.64898/2026.06.09.26355167

**Authors:** Heesoo Joo, Brian Maskery, Alexander Klosovsky, Marlon Marques, Salma Taher, Warren Dalal, Michelle Weinberg, Tarissa Mitchell

**Author notes:** Corresponding Author: Brian Maskery, PhD, Division of Global Migration Health, US Centers for Disease Control and Prevention 1600 Clifton Road, MS H16-4, Atlanta, GA 30329, USA. Brian Maskery and Tarissa Mitchell contributed equally as senior authors. The findings and conclusions in this report are those of the authors and do not necessarily represent the official position of the Centers for Disease Control and Prevention or the institutions with which the authors are affiliated.

## Abstract

**Context:** To ensure healthy resettlement and protect US health security, the Vaccination Program for US-bound Refugees (VPR) offers some recommended vaccines to refugees overseas before resettlement to the United States. The selected vaccines and number of doses vary by country of departure. VPR was found to be cost-saving in 2018 but had since expanded to more sites.

**Objective:** Assess VPR’s current costs and impact on post-arrival domestic vaccination needs and costs.

**Setting and Participants:** A model-based analysis of the Federal government costs for VPR and post-arrival (US) vaccination of resettled refugees separated across five regions: Africa, Asia, the Middle East and North Africa/Republic of Türkiye and Middle East, Europe, and the Americas using fiscal year 2024 data.

**Design:** We quantified and compared full vaccination costs for refugees under two scenarios: (1) ‘No VPR’ and (2) ‘VPR’. Refugees would receive no vaccines overseas and be fully vaccinated after US arrival under ‘No VPR’. Under ‘VPR’, refugees receive one or two doses of selected vaccines overseas before completing vaccination schedules after arrival.

**Main Outcomes:** Costs were reported in 2023 US dollars for ‘VPR’ and ‘No VPR’ scenarios and further subdivided by grouping countries/sites depending on whether the International Organization for Migration (IOM) provides vaccination services for refugees (IOM sites) versus non-IOM providers (non-IOM sites).

**Results:** ‘VPR’ resulted in average net cost savings of $147 per person or $14.7 million per 100,000-refugee cohort compared to providing all vaccines after US arrival (‘No VPR’). ’VPR’ was cost-saving across most regions, except for IOM sites in Europe, where a net cost of $44 per person was observed. Net cost savings per person were highest for IOM sites in Africa ($333).

**Conclusions:** VPR remains a cost-saving strategy, while protecting US-bound refugees’ health and US health security by preventing disease outbreaks during resettlement.

**Implications for Policy & Practice:**

- The Vaccination Program for US-bound Refugees (VPR) provides some vaccines to refugees overseas, before they depart for the United States.
- VPR reduces costs relative to vaccination after US arrival.
- Net cost savings with VPR are $14.7 million per 100,000-refugee cohort and vary from - $44 to $333 per person, depending on the region.
- Vaccination prior to departure reduces the likelihood that refugees would experience illness or costly delays during travel, or arrive while infected with vaccine-preventable diseases such as measles or polio, which may impose additional response costs to mitigate transmission inside the United States.
- Other countries may consider this analysis of potential net cost savings when considering the use of pre-departure vaccination as part of their refugee resettlement program.

## 1. Introduction

In response to health security risks during refugee resettlement, including the importation of vaccine-preventable diseases (VPD), the US Centers for Disease Control and Prevention (CDC) and the US Department of State established the overseas Vaccination Program for US-bound Refugees (VPR) in 2012 ^1^. Initially piloted in two countries in December 2012 ^1^, this voluntary program expanded to over 80 countries processing US-bound refugees by 2024. The International Organization for Migration (IOM) typically conducts overseas health assessments for US-bound refugees. However, recently, new countries where IOM does not conduct overseas medical processes have been added. For example, in fiscal year (FY) 2024, (i.e., October 2023 to September 2024) over 30% of overseas health assessments were conducted in such countries.

These panel sites, operated by trained and licensed medical doctors appointed by the local US embassy or consulate, perform required medical examinations for US residency applicants, including US-bound refugees ^2^. Unlike immigrants, refugees are not required to receive vaccines during overseas medical examinations, although vaccination is mandatory at the time of adjustment of status to lawful permanent resident, which they can apply for one year after US arrival.

The VPR was previously evaluated in 2018 and found effective in reducing US government costs ^3^. Compared to the baseline scenario where all vaccinations would be administered post-arrival, estimated net cost savings per US-bound refugee in 2018 ranged from $225.93 to $498.42 in 2015 US dollars ^3^.

In 2024, the full VPR immunization schedule included 12 vaccines with up to two doses per vaccine series offered to refugees before travel, depending on age and past vaccination history. This allowed refugees to start the US-recommended immunization schedule before resettlement (with some differences related to disease risk, vaccine cost, or availability). Due to cost and logistical constraints, non-IOM panel sites followed an abbreviated VPR schedule, which in FY2024 included seven vaccines, administering one dose of each age-appropriate vaccine. All US-bound refugees were offered the hepatitis B surface antigen (HBsAg) test to determine infection status before vaccination. IOM and non-IOM panel sites document all VPR-administered vaccines and pre-VPR valid, documented vaccines in an official immunization record, shared with state health departments at refugees’ final destinations upon US arrival.

The 2018 analysis used FY2017 program budget data, which included fewer vaccines and departure countries. Our objective was to update this analysis with FY2024 budget data and departure countries to identify regional cost differences and the impact of adding non-IOM sites. To align with FY2024 budget data, this analysis used the CDC recommended Immunization Schedule as of calendar year 2024.We performed a comparative cost analysis of completing full vaccination schedules after US arrival versus the combination of VPR-administered doses overseas and US follow-up for any remaining doses.

## 2. Methods

We compared costs for US-bound refugees to complete age-specific vaccination schedules under two scenarios: (1) ‘No VPR’, and (2) ‘VPR’ using methods adapted from Joo et al. (2018) ^3^. For ‘No VPR’, refugees receive all vaccines domestically after US arrival. For ‘VPR’, they receive one or two doses of each vaccine overseas before departure and complete their vaccination schedules after arrival. All costs were estimated in 2023 US dollars from the US payers’ perspective ^4^ using a one-year time horizon.

We included age-specific, 2024 CDC-recommended vaccines except COVID-19 and seasonal influenza ^5–7^. COVID-19 vaccines were excluded because US-bound refugees were referred to national programs. The seasonal influenza vaccine was excluded due to its limited use pre-arrival. The FY2024 VPR schedule recommended one to two doses of the following vaccines overseas, depending on age and vaccination history (Appendix Tables A1-A3): diphtheria, tetanus, and pertussis (DTP); tetanus and diphtheria (Td); hepatitis B adult (HepB) and pediatric (HepB Ped); pentavalent (HepB-*Haemophilus influenzae* type b (Hib)-DTP); inactivated poliovirus (IPV) or bivalent oral poliovirus (bOPV) where IPV was unavailable; measles, mumps, and rubella (MMR); rotavirus; meningococcal conjugate (MenACWY); pneumococcal conjugate (PCV); and varicella (VAR). Vaccine availability and number of doses offered through VPR varied by country. US-bound refugees were categorized into five groups based on their departure regions: Africa, Asia, the Middle East and North Africa (MENA)/Republic of Türkiye and Middle East (TUME), Europe and the Americas. We also considered whether vaccination was conducted by IOM or non-IOM panel sites in each country, resulting in nine groups; the Americas had no IOM panel sites during our analysis period.

### Domestic vaccination costs for US-bound refugees

Domestic (i.e., US) vaccination costs encompassed both vaccine costs and administration fees (Appendix Tables A4-A7). In FY 2024,,most US-bound refugees were covered under Medicaid or the Refugee Medical Assistance (RMA) program for up to 12 months after arrival ^8^. Given the one-year time horizon, we assumed most vaccinations would be covered by Medicaid or RMA. RMA reimbursement rates aligned closely with Medicaid rates, and the Vaccines for Children (VFC) program provides vaccines to Medicaid beneficiaries aged 0 to 18 years ^9^.

Baseline and lower-bound cost estimates for pediatric vaccines were derived from the *Pediatric/VFC Vaccine Price List* as of July 1, 2023 (CDC costs); the same list’s private sector costs were used for upper-bound estimates ^10^.

Baseline costs for adult vaccines were estimated using 2021 MarketScan Medicaid multi-state data, adjusted to 2023 prices using average changes in private sector costs between 2021 and 2023 from the CDC *Adult Vaccine Price List* as of July 1, 2023 ^10^. Lower- and upper-bound estimates were drawn from CDC and private sector costs from the same list, respectively ^10^.

We estimated baseline and lower-bound vaccine administration fees for Medicaid beneficiaries using Current Procedural Terminology (CPT) codes 90460 and 90461 for children, and CPT codes 90471 and 90472 for adults following Joo et al. (2018) ^3^. The upper-bound cost estimate was derived from the 2023 Medicare Physician Fee Schedule ^11^.

### Overseas vaccination costs for US-bound refugees

To estimate overseas vaccine and administration costs, we used FY2024 country-specific vaccination budgets (Appendix Tables A8-A10). We calculated the weighted average cost per dose by vaccine and region, based on the expected number of US-bound refugees and anticipated vaccine purchases. For program administration costs, we assumed some cost components were incurred on a per-dose basis while others were incurred per-capita. Per-dose costs included operational costs (excluding vaccine-specific prices) and administration fees for non-IOM sites. Per-capita costs, independent, of numbers of doses administered, included expenses for office space, staffing, cold chain maintenance, and HBsAg testing. We applied a 7% overhead multiplier to all estimates and assumed per-dose or per-capita costs from the FY2024 budget were equivalent to 2023 costs.

We used FY2024 medical caseloads of 72,700 refugees undergoing health assessments at IOM sites and 104,700 at non-IOM sites. These caseloads included some individuals undergoing multiple overseas health assessments, during which they may have received additional vaccine doses but not HBsAg retesting. US-bound refugees require another health assessment (referred to as re-medicals) if they do not depart within a defined period after their initial assessments.

Therefore, the number of overseas health assessments (medical caseload) exceeded the number of refugee arrivals. Budget estimates were pro-rated to account for individuals receiving fewer doses due to re-medicals, as fewer doses are needed at subsequent health assessment. We present cost estimates for each scenario per refugee and for a cohort of 100,000 refugees, based on the projected distribution of departures across regions and IOM versus non-IOM sites for the FY2024 budget. The 100,000-refugee cohort approximates the number of refugees resettled into the United States during FY2024.

### One-way sensitivity analysis

We conducted one-way sensitivity analyses to evaluate the robustness of the baseline analysis by incorporating a range of domestic vaccination costs using the lower- and upper-bound estimates defined above.

### Sub-model analyses

We conducted three sub-model analyses to address uncertainties related to VPR implementation overseas, focusing on key factors influencing costs under the ‘VPR’ scenario: (1) Sub-model 1: Historical vaccine records documentation, which incorporates cost savings from reduced domestic doses due to availability of overseas historical vaccine records; (2) Sub-model 2: Alternative vaccine cost–estimation based on potentially lower vaccine prices for the Americas, and (3) Sub-model 3: bOPV utilization and IPV re-vaccination, accounting for overseas bOPV use, which necessitates domestic IPV re-vaccination costs.

### Impact of VPR on VPD outbreaks

Prevention of US VPD outbreaks by reducing importation is another key VPR benefit. We estimated costs associated with overseas measles and polio outbreaks among US-bound refugees, including US domestic response efforts if a case is imported. While VPR mitigates this risk, we accounted for these potential cost savings separately from the main (‘VPR’ versus “No VPR’) analysis.

Further details on the data and methods used for the analysis are provided in the Appendix. This activity was reviewed by CDC, deemed not research, and conducted consistent with applicable federal law and CDC policy.^§^ Existing text in this manuscript was edited using the CDC chatbot based on ChatGPT-5-mini (OpenAI, San Francisco CA) to improve conciseness.

## 3. Results

The average full vaccination cost was $1,102.7 per person for the ‘No VPR’ scenario and $955.7 per person for ‘VPR’, resulting in net cost savings of $147.0 per person (Table 1). Total estimated costs per person were $872.2 for IOM sites and $1,013.7 for non-IOM sites under ‘VPR’; average estimated costs of overseas vaccination per person were $52.2 for IOM sites and $54.4 for non-IOM sites. Average costs to complete remaining age-appropriate 2024 CDC-recommended vaccination schedules in the United States were estimated at $820.0 for refugees from IOM sites and $959.3 for those from non-IOM sites. By comparison, if all vaccinations were administered domestically after arrival under ‘No VPR’, total estimated costs per person would be $1,136.9 for IOM sites and $1,079.0 for non-IOM sites.

**Table 1:** Costs of full vaccination per US-bound refugee, excluding influenza and COVID-19 vaccines, with and without the VPR, by region (2023 US dollars).

|  | Region | 'No VPR' scenario, (A) | 'VPR' scenario |  |  | Net cost savings, (E)= (A)–(B) | Percent cost reduction, (E)/(A) | Total number of doses*, (F) | Number of overseas VPR doses, (G) | Net cost saving per overseas VPR dose, (E)/(G) |
| --- | --- | --- | --- | --- | --- | --- | --- | --- | --- | --- |
|  |  |  | Total, (B)=(C)+(D) | Overseas, (C) | United States, (D) |  |  |  |  |  |
| IOM | Africa | \$1,175.6 | \$843.1 | \$40.3 | \$802.8 | \$332.5 | 28% | 12.8 | 5.3 | \$63.3 |
| | Asia | \$1,137.6 | \$887.1 | \$88.0 | \$799.1 | \$250.5 | 22% | 11.8 | 4.8 | \$51.8 |
| | MENA/TUME | \$1,086.7 | \$877.8 | \$41.4 | \$836.5 | \$208.8 | 19% | 11.8 | 4.0 | \$52.1 |
| | Europe | \$1,045.7 | \$1,089.8 | \$86.2 | \$1,003.6 | –\$44.1<sup>†</sup> | –4%<sup>†</sup> | 11.4 | 0.6 | –\$77.8<sup>†</sup> |
| | Total | \$1,136.9 | \$872.2 | \$52.2 | \$820.0 | \$264.7 | 23% | 12.5 | 4.6 | \$57.3 |
| Non–IOM | Africa | \$1,148.6 | \$965.9 | \$34.0 | \$931.9 | \$182.8 | 16% | 12.6 | 3.6 | \$50.2 |
| | Asia | \$1,099.5 | \$954.1 | \$53.7 | \$900.4 | \$145.4 | 13% | 11.7 | 2.5 | \$58.3 |
| | MENA/TUME | \$1,067.2 | \$971.9 | \$33.1 | \$938.8 | \$95.3 | 9% | 11.8 | 2.1 | \$46.1 |
| | Europe | \$1,024.8 | \$1,004.8 | \$63.8 | \$941.1 | \$20.0 | 2% | 11.1 | 1.1 | \$17.8 |
| | Americas | \$1,067.9 | \$1,046.5 | \$62.7 | \$983.9 | \$21.3 | 2% | 11.9 | 1.2 | \$17.3 |
| | Total | \$1,079.0 | \$1,013.7 | \$54.4 | \$959.3 | \$65.3 | 6% | 12.1 | 1.8 | \$37.0 |
| Grand Total | | \$1,102.7 | \$955.7 | \$53.5 | \$902.2 | \$147.0 | 13% | 12.2 | 2.9 | \$50.1 |

Abbreviations: Coronavirus Disease 2019 (COVID-19); Vaccination Program for US-bound Refugees (VPR); Middle East and North Africa (MENA); the Republic of Türkiye and Middle East (TUME); International Organization for Migration (IOM)
Notes: \*The total number of doses was estimated based on age distribution in each region. The variation in doses among regions is attributed to differences in the age distribution of refugees.
<sup>†</sup>The '–' signs represent net cost or cost increase

Overall, vaccination costs for ‘VPR’ were lower than those for ‘No VPR’ in eight out of nine regions (Table 1). The only exception was for IOM panel sites in Europe, where the average full vaccination cost per person under ‘No VPR’ scenario was $1,045.7 compared to $1,089.8 under ‘VPR’ (net cost: $44.1 per person). The highest net cost savings were estimated for IOM panel sites in Africa (net cost savings: $332.5 per person).

Excluding seasonal influenza and COVID-19 vaccines, US-bound refugees examined in IOM panel sites were recommended to receive 12.5 doses of vaccines on average per person while those examined in non-IOM sites were recommended to receive 12.1 doses on average, based on the 2024 CDC-recommended immunization schedule(Table 1 and Appendix Tables A2-A3). The slight variation is attributed to differences in age distributions. Overseas, the VPR was projected to provide an average of 4.6 doses per person overseas in IOM sites (7.9 subsequent doses needed post-arrival) and 1.8 doses per person in non-IOM sites (10.3 doses post-arrival).

On average (across all sites), 2.9 doses per person were projected to be administered overseas, yielding net cost savings of $50.1 per overseas dose (Table 1). The highest number of overseas doses per person were projected for US-bound refugees departing from IOM sites in Africa, Asia, and MENA/TUME (range: 4.0 to 5.3) with net cost savings per overseas dose ranging from $51.8 to $63.3 and net cost savings per person from $208.8 to $332.5. In contrast, refugees from non-IOM sites in the same regions received between 2.1 to 3.6 doses, with net cost savings per person ranging from $95.3 to $182.8. The net cost savings per overseas dose estimates were similar for non-IOM and IOM sites in Africa, Asia, and MENA/TUME (range: $46.1 to $63.3). The projected numbers of overseas doses per person were much lower for US-bound refugees departing from non-IOM sites in Europe and the Americas (range: 1.1 to 1.2), with net cost savings per overseas dose of only $17.3 to $17.7 and net cost savings per person of $20.0 to $21.3.

The overseas vaccination cost per 100,000-refugee cohort was estimated at $5.4 million and the domestic US vaccination cost at $90.2 million ($95.6 million total) under the ‘VPR’ scenario (Table 2). Under the ‘No VPR’ scenario, administering the same vaccine doses domestically would cost an estimated $110.3 million. Thus, relative to ‘No VPR’, ‘VPR’ was projected to save $14.7 million per 100,000-refugee cohort, including $10.8 million for US-bound refugees from IOM sites and $3.9 million from non-IOM sites. The largest cost savings occurred in IOM sites in Africa ($6.4 million, 43% of total net cost savings). Only IOM sites in Europe had a net cost increase of $75,000 with ‘VPR’ compared to ‘no VPR’.

**Table 2:** Total costs of full vaccination of US-bound refugees with and without the VPR per 100,000-refugee cohort, excluding influenza and COVID-19 vaccines, by region (2023 million US dollars).

|  | Region | No VPR' scenario, (A) | VPR' scenario |  |  | Net cost savings, (E)=(A)-(B) | Medical caseload per 100,000 refugee cohort |
| --- | --- | --- | --- | --- | --- | --- | --- |
|  |  |  | Total, (B)=(C)+(D) | Overseas, (C) | United States, (D) |  |  |
| IOM | Africa | \$22.5 | \$16.2 | \$0.8 | \$15.4 | \$6.4 | 19,166 |
| | Asia | \$9.4 | \$7.4 | \$0.7 | \$6.6 | \$2.1 | 8,286 |
| | MENA/TUME | \$12.9 | \$10.4 | \$0.5 | \$9.9 | \$2.5 | 11,838 |
| | Europe | \$1.8 | \$1.8 | \$0.1 | \$1.7 | – \$0.1 | 1,691 |
|  | Americas | N/A | N/A | N/A | N/A | N/A | N/A |
| | Total | \$46.6 | \$35.7 | \$2.1 | \$33.6 | \$10.8 | 40,981 |
| Non-IOM | Africa | \$6.4 | \$5.4 | \$0.2 | \$5.2 | \$1.0 | 5,581 |
| | Asia | \$9.5 | \$8.2 | \$0.5 | \$7.8 | \$1.3 | 8,625 |
| | MENA/TUME | \$9.0 | \$8.2 | \$0.3 | \$7.9 | \$0.8 | 8,455 |
| | Europe | \$1.4 | \$1.4 | \$0.1 | \$1.3 | \$0.0 | 1,409 |
| | Americas | \$37.3 | \$36.6 | \$2.2 | \$34.4 | \$0.7 | 34,949 |
| | Total | \$63.7 | \$59.8 | \$3.2 | \$56.6 | \$3.9 | 59,019 |
| Grand Total | | \$110.3 | \$95.6 | \$5.4 | \$90.2 | \$14.7 | 100,000 |

Abbreviations: Vaccination Program for US-bound Refugees (VPR); Middle East and North Africa (MENA); the Republic of Türkiye and Middle East (TUME); International Organization for Migration (IOM)
Notes: \* The ‘–’ signs represent net cost or cost increase. The 100,000-refugee cohort is weighted by population using the 177,400 medical caseload included in the FY2024 budget. Each refugee is assumed to undergo one overseas health assessment. Some refugees will need one or more health assessments (referred to re-medicals) at which time they would be eligible to receive additional vaccine doses. The costs and number of vaccine doses received during re-medicals are not considered in this analysis.

The one-way sensitivity analysis of domestic vaccination prices indicated that total net cost savings per 100,000-refugee cohort could have ranged from $11.5 million to $24.6 million, compared to the baseline estimate of $14.7 million. Further details are provided in Appendix Table A11.

As detailed in Appendix Tables A15-A19, the estimated impact of VPR on reducing the risk of VPD outbreaks would yield additional annual cost savings of $1.2 million (range: $0.6–$10.1 million). Without the VPR, measles outbreaks could incur an additional $0.5 million (range: $0.3–$9.0 million) annually in costs associated with pausing travel, implementing outbreak recommendations for US-bound refugees, or managing imported cases. Similarly, polio outbreaks, though less frequent than measles outbreaks, could lead to additional costs of $0.7 million (range: $0.3 million–$1.2 million) per year without the VPR, based on costs for re-vaccination and post-arrival domestic testing associated with a 2006 polio outbreak ^12^.

## 4. Discussion

VPR remained a cost-saving strategy in 2024, with average net cost savings of $147.0 per person. This finding aligns with previous analyses, which estimated net cost savings of $226 per person in 2015 US dollars ($290 per person in 2023 US dollars) ^3^, and net cost savings of $235 per person in 2005 US dollars ($367 per person in 2023 US dollars) ^13^.

The reduction in net cost savings per person in the current analysis compared to the 2018 evaluation is primarily due to shifts in the regional distribution of US-bound refugees and variation in vaccination costs and doses delivered by region. In 2018, few US-bound refugees were processed in non-IOM panel sites. In contrast, in 2024 approximately 35% of the annual 100,000-refugee cohort was examined in non-IOM panel sites in the Americas. Further, the number of vaccine doses provided to US-bound refugees in high-cost regions, such as Europe and the Americas (range: 0.6 to 1.2 overseas VPR doses from Table 1), is less than 2018 estimates (an average of 6.4 overseas VPR doses from Appendix Table A24). Since US-bound refugees departing from the Americas receive fewer doses, and the cost per dose delivered through VPR is higher, net cost savings per person from the Americas were only $21.3 for 1.2 overseas doses, compared to an average of $264.7 for 4.6 overseas doses per person from IOM sites outside the Americas. Excluding refugees resettling from the Americas, net cost savings were estimated at $214.5 per person, which is 46% greater than the baseline estimate of $147 (Appendix Table A24). Efforts to reduce vaccination costs in the Americas, such as those proposed in Sub-model 2, could further improve net cost savings from VPR.

Net cost savings per overseas VPR dose were achieved in three out of four regions for IOM sites and all five regions for non-IOM sites. Providing additional vaccine doses through VPR (e.g., second doses of VPR-recommended vaccines across all sites) should yield greater net cost savings for the US government compared to domestic administration. However, the impact varies by region. Adding doses in Africa, Asia, and MENA/TUME regions would likely generate greater savings compared to the Americas or Europe. Conversely, providing additional doses through VPR in European IOM sites may decrease cost savings. However, overseas VPR costs European IOM sites are likely overestimated because IOM staff in these countries oversaw VPR activities at non-IOM sites in Europe and included these support costs within their 2024 vaccination budget. As only 3.1% of the 2024 medical caseload was from Europe, higher costs in Europe are unlikely to significantly impact overall VPR costs, if caseloads remain low. Additionally, we did not consider VPR’s impact on VPD outbreak prevention in the main analysis, resulting in underestimated costs for the ‘No VPR’ scenario. Including reduced VPD outbreak response costs could result in net cost-savings for VPR implementation at IOM sites in Europe, especially given the increasing incidence of measles in the region ^14^. Still, further shifts in refugee health assessments from Africa, Asia, and MENA/TUME to Europe or the Americas could increase costs and reduce net cost savings for the ‘VPR’ scenario.

Our baseline analysis provided a conservative estimate of VPR’s net cost savings of $14.7 million per 100,000-refugee cohort, assuming that RMA or Medicaid would cover all domestic vaccination costs for refugees for up to one year after arrival in FY 2024. Some refugees may obtain private insurance or pay out of pocket, leading to higher payments for U.S. doses; the upper-bound annual ‘VPR’ net cost savings from one-way sensitivity analyses was $24.6 million relative to ‘No VPR’ using private sector prices. Across three Sub-models, the best-case scenario combines Sub-models 1 and 2, while the worst-case scenario is represented by Sub-model 3 alone. The range of net cost savings across the three Sub-models varies from $14.5 to $20.0 million per 100,000-refugee cohort, suggesting that net cost savings may be underestimated or increased through changes in vaccine procurement.

This evaluation has limitations. First, we did not account for potential net cost savings from preventing VPD outbreaks beyond polio and measles. For example, although diphtheria and pertussis vaccines are available through VPR, we did not include outbreak response costs due to limited cost data for these VPD outbreaks among refugees. Similar to polio and measles, overseas diphtheria or pertussis outbreaks could increase refugee resettlement costs and US response costs. Therefore, the $1.2 million in estimated net cost savings from VPD outbreak prevention is likely underestimated.

Next, some healthcare providers may not use overseas vaccination records generated through VPR when providing vaccines to refugees after arrival, potentially leading to overestimation of VPR’s net cost savings. However, an earlier analysis showed that VPR reduced the number of vaccines delivered at refugees’ initial post-arrival domestic medical assessments ^15^. This analysis was conducted shortly after VPR was initiated. The ongoing operation of VPR and state refugee health partners’ familiarity with VPR should mitigate the risk of unnecessary re-vaccination. Further, vaccination records from overseas refugee health assessments are increasingly transferred directly into US state immunization information systems through the Refugee Immunization Information Systems Exchange (RIISE) project, facilitating access for US medical providers.^16^

Finally, our analysis was based on budget estimates of the projected number of vaccines and costs for VPR. Actual doses delivered and costs may deviate from these projections. Since VPR has operated for more than 10 years, we believe budget estimates are likely close to actual costs and doses delivered per person during a typical year.

If some individuals who underwent overseas refugee health assessments did not ultimately resettle to the United States, our per-person net cost savings estimates with VPR may be overestimated. Conversely, if some refugee health assessments were conducted in FY2024 for refugees arriving in FY2025, these estimates would be unaffected (i.e., savings would simply be delayed until the next fiscal year). Total net cost-savings depend on both medical caseloads (i.e., number of overseas refugee health assessments) and refugee arrivals. In FY 2024, there were 100,000 US-bound refugee arrivals compared to a 177,000-person medical caseload, which included individuals requiring re-medicals. The budgeted cost accounts for the reduced doses received per person due to re-medicals but does not include separate operational cost estimates for vaccine doses delivered at initial overseas health assessments versus re-medicals. We assumed that operational costs per dose during re-medicals were similar to costs for initial health assessments when presenting results per 100,000-refugee cohort.

## Conclusions

Our findings align with previous analyses, demonstrating that VPR reduces vaccination costs for US-bound refugees compared to administering all vaccines post US arrival. We estimated VPR-associated annual net cost savings of approximately $14.7 million for a cohort of 100,000 resettled refugees. Expanding VPR to cover additional vaccines would likely increase the US federal government’s net cost savings; net cost savings may decrease if the fraction of refugees resettled from Europe or the Americas increases.

## Footnote

§See e.g., 45 C.F.R. part 46, 21 C.F.R. part 56; 42 U.S.C. §241(d); 5 U.S.C. §552a; 44 U.S.C.

§3501 et seq.

## Data availability

The data used for this analysis are accessible from a third-party source, Merative®. Data availability is subject to restrictions as the data were utilized under license and are not publicly accessible.

## List of Supplemental Digital Content

All supplemental content is included in an appendix submitted with this manuscript.

## Supporting information

Supplemental Digitial Content (Appendix)

## Data Availability

The data used for this analysis are accessible from a third-party source, Merative(R). Data availability is subject to restrictions as the data were utilized under license and are not publicly accessible.

