## Supplemental Digitial Content (Appendix) for "Cost analysis of overseas versus domestic vaccination of US-bound refugees"

### 1. Overview

After a decade of responding to health security risks during refugee resettlement, the US Centers for Disease Control and Prevention (CDC) and the US Department of State developed the Vaccination Program for US-bound Refugees (VPR) [1]. This voluntary, overseas immunization program was first piloted in two countries in December 2012 [1] and is now offered in all countries (>80) processing refugees. The main implementing partner is the International Organization for Migration (IOM). Vaccines are delivered by panel physicians in the countries from which refugees depart. IOM directly employs panel physicians in some countries to support refugee resettlement. However, refugee health assessments are being conducted by non-IOM panel physicians in other countries where IOM does not directly conduct refugee health assessments. We separately consider the costs and impact of VPR depending on whether IOM directly supports refugee health assessments by country. Thus, we group the countries in each region as IOM sites and non-IOM sites. The number of non-IOM sites has increased in recent years as the number of countries from which US-bound refugees has increased.

In 2018, we quantified and compared the full vaccination costs for refugees using two scenarios: (1) a comparator of no VPR and (2) the current situation (2018) with VPR [2]. At the time of evaluation, we used fiscal year (FY) 2017 budget data, including 20 countries with eight vaccines (diphtheria, tetanus, and pertussis (DTP); tetanus and diphtheria (Td); hepatitis B adult (HepB) and pediatric (HepB Ped); Pentavalent (HepB-*Haemophilus* influenzae type b (Hib)-DTP); inactivated poliovirus (IPV) or oral poliovirus (OPV); measles, mumps, and rubella (MMR); pneumococcal conjugate (PCV)); and Hib) for the overseas vaccination program [2]. With scenario 1 (no VPR), all refugees started ACIP recommended vaccinations after their resettlement in the United States. With scenario 2 (VPR), we assumed that all refugees received two doses of up to eight vaccines based on their age prior to departure through VPR and completed the series and other US Advisory Committee on Immunization Practices

(ACIP) recommended vaccines in the United States. We found that the overseas vaccination program was a cost-saving strategy that also reduced the risk of refugees arriving while infected with a vaccine-preventable disease. Vaccination costs with the (2) VPR scenario were lower than costs for (1) no VPR scenario for refugees in all examined age groups. Relative to (1) no VPR, Net cost savings per person associated with (2) VPR ranged from $225.93 with estimated Refugee Medical Assistance (RMA) or Medicaid payments for domestic costs to $498.42 with estimated private sector payments in 2015 US dollars [2]. Net cost savings with the (2) VPR scenario relative to (1) no VPR scenario were sensitive to inputs for vaccination costs, domestic vaccine coverage rates, and revaccination rates, but (2) VPR scenario was cost-saving across a range of plausible parameter estimates. The 2018 analysis did not account for differences between IOM and non-IOM sites.

In 2024, the full VPR program features an immunization schedule of up to 12 vaccines (i.e., DTP; Td; HepB and HepB Ped; Pentavalent; IPV or bivariant oral poliovirus (bOPV); MMR; rotavirus; meningococcal conjugate (MenACWY); PCV; and varicella (VAR)) in more than 80 countries. Refugees are offered up to two doses of one or more age-appropriate vaccines before departure. Due to cost and logistical considerations, non-IOM panel sites follow an abbreviated immunization schedule of only seven vaccines, offering one dose of each age-appropriate vaccine. In addition, the number of vaccines provided in each country may vary depending on cost and availability. We conducted an analysis of the proposed FY2024 budget to identify differences in the costs of providing overseas, pre-departure vaccine doses to US-bound refugees by region, depending on whether vaccination is provided or facilitated by IOM versus non-IOM panel sites. Additionally, we conducted a comparative cost analysis of providing the same vaccine doses after arrival in the United States.

### 2. Decision Tree

We adapted the decision tree model used in Joo et al. (2017), which was originally stratified by age [2]. In this study, the model was stratified by region and whether a country has IOM panel sites. The regions considered were Africa, Asia, Middle East and North Africa (MENA)/Türkiye and Middle East (TUMA), Europe, and America. These stratifications were selected because of significant variations in vaccination costs across these factors. Notably, in contrast to the 2018 analysis, the types of vaccines provided through VPR varied across countries based on data used to develop the VPR budget.

We analyzed and compared the costs for refugees to complete the age-specific vaccination schedule recommended by the ACIP under two scenarios: (1) without VPR, referred to as the 'No VPR' scenario, and (2) with VPR, referred to as the 'VPR' scenario. Again, these two scenarios were adapted from Joo et al. (2017) [2]. In the 'VPR' scenario, US-bound refugees received one or two doses of each vaccine provided by VPR as projected in country-specific budgets before departure and complete their age-specific vaccine schedules after resettlement in the United States. In the 'No VPR' scenario, refugees receive all vaccines after resettlement in the United States, completing the ACIP-recommended schedule at that time.

### 3. The number of vaccine doses

All ACIP-recommended vaccines, excepts influenza and Coronavirus Disease 2019 (COVID-19) vaccines, were included in the analyses (Table A1). While all these vaccines can be administered in the United States, VPR is unable to provide certain ACIP-recommended vaccines overseas. Table A1 lists all vaccines included in the FY 2024 IOM budget for VPR, excluding influenza and COVID-19 vaccines. However, the types of vaccines provided by VPR vary by country.

Table A1. List of examined vaccines and the availability of each vaccine in the United States and overseas.

| Vaccine | Overseas (VPR) | United States |
|---|---|---|
| Diphtheria, tetanus, and pertussis vaccine (DTP) | o | o |

| | | |
|---|---|---|
| Tetanus, diphtheria, and acellular pertussis vaccine (Tdap) | × | o |
| Tetanus, and diphtheria vaccine (Td) | o | o |
| Hepatitis B adult vaccine (HepB) | o | o |
| Hepatitis B pediatric vaccine (HepB Ped) | o | o |
| Pentavalent vaccine including diphtheria, tetanus, pertussis, *Haemophilus influenzae* type b (Hib), and hepatitis B antigens (Penta) | o | o |
| Inactivated poliovirus vaccine (IPV) or Bivalent oral poliovirus vaccine (bOPV)[1] | o | o |
| Measles, mumps, and rubella vaccine (MMR) | o | o |
| Rotavirus vaccine (Rota) | o | o |
| Meningococcal conjugate vaccine (MenACWY) | o | o |
| Pneumococcal conjugate vaccine (PCV) | o | o |
| Varicella vaccine (VAR) | o | o |
| Hepatitis A vaccine (HepA) | × | o |
| Human papillomavirus vaccine (HPV) | × | o |

[1] In the United States, bOPV is not recommended by ACIP. However, VPR cannot provide IPV in some countries and provides bOPV instead.

The numbers of vaccine doses recommended by the ACIP varies based on age [3-5]. To account for this, we calculated the weighted average number of doses for each vaccine, considering the age distribution of the US-bound refugees within country (Tables A2-A3). Since age distribution differs by region and for IOM versus non-IOM sites, the total weighted average number of doses also varies by region and IOM/non-IOM site. For instance, regions with higher proportions of adult refugees would have lower weighted average numbers of recommended doses compared to regions with higher proportions of child refugees. The number of doses delivered overseas for the 'VPR' scenario is based on the projected number of doses delivered according to FY24 IOM budget data.

Table A2: The number of doses for completing full vaccination by region for IOM sites

**'No VPR' scenario**

| Vaccination location | Region | DTP | Tdap /Td | HepB | HepB Ped | Penta | IPV/ bOPV | MMR | Rota | MenACWY | PCV | VAR | HepA | HPV | Total |
|---|---|---|---|---|---|---|---|---|---|---|---|---|---|---|---|
| US | Africa | 0.51 | 2.40 | 1.31 | 1.43 | 0.26 | 0.37 | 1.94 | 0.04 | 0.39 | 0.52 | 1.61 | 1.13 | 1.11 | 13.02 |
| | Asia | 0.53 | 2.29 | 1.68 | 0.98 | 0.34 | 0.18 | 1.92 | 0.05 | 0.22 | 0.68 | 1.50 | 0.88 | 0.91 | 12.16 |
| | MENA/TUME | 0.45 | 2.44 | 1.64 | 1.14 | 0.22 | 0.33 | 1.82 | 0.06 | 0.24 | 0.44 | 1.51 | 0.91 | 0.87 | 12.06 |
| | Europe | 0.27 | 2.62 | 1.83 | 1.02 | 0.15 | 0.16 | 1.83 | 0.06 | 0.32 | 0.30 | 1.45 | 0.78 | 0.75 | 11.56 |
| | Total | 0.48 | 2.40 | 1.50 | 1.24 | 0.26 | 0.31 | 1.90 | 0.05 | 0.31 | 0.52 | 1.55 | 1.00 | 0.99 | 12.51 |

**'VPR' scenario**

| Vaccination location | Region | DTP | Tdap /Td | HepB | HepB Ped | Penta | IPV/ OPV | MMR | Rota | MenACWY | PCV | VAR | HepA | HPV | Total |
|---|---|---|---|---|---|---|---|---|---|---|---|---|---|---|---|
| Overseas | Africa | 0.06 | 1.13 | 0.65 | 0.80 | 0.20 | 0.17 | 1.46 | 0.03 | 0.02 | 0.18 | 0.56 | N/A | N/A | 5.26 |
| | Asia | 0.03 | 1.15 | 0.84 | 0.66 | 0.25 | 0.06 | 1.44 | 0.03 | N/A | 0.00 | 0.37 | N/A | N/A | 4.83 |
| | MENA/TUME | 0.05 | 0.83 | 0.82 | 0.61 | 0.15 | 0.17 | 1.36 | N/A | N/A | 0.01 | N/A | N/A | N/A | 4.01 |
| | Europe | 0.02 | N/A | N/A | N/A | 0.04 | 0.03 | 0.47 | N/A | N/A | N/A | N/A | N/A | N/A | 0.57 |
| | Total | 0.05 | 1.00 | 0.71 | 0.68 | 0.19 | 0.14 | 1.39 | 0.02 | 0.01 | 0.09 | 0.34 | N/A | N/A | 4.62 |
| US | Africa | 0.45 | 1.27 | 0.65 | 0.63 | 0.07 | 0.19 | 0.49 | 0.01 | 0.37 | 0.34 | 1.06 | 1.13 | 1.11 | 7.76 |
| | Asia | 0.50 | 1.15 | 0.84 | 0.32 | 0.08 | 0.11 | 0.48 | 0.03 | 0.22 | 0.68 | 1.12 | 0.88 | 0.91 | 7.32 |
| | MENA/TUME | 0.40 | 1.60 | 0.82 | 0.53 | 0.07 | 0.17 | 0.45 | 0.06 | 0.24 | 0.43 | 1.51 | 0.91 | 0.87 | 8.05 |
| | Europe | 0.25 | 2.62 | 1.83 | 1.02 | 0.11 | 0.13 | 1.36 | 0.06 | 0.32 | 0.30 | 1.45 | 0.78 | 0.75 | 10.99 |
| | Total | 0.44 | 1.40 | 0.79 | 0.56 | 0.07 | 0.17 | 0.51 | 0.03 | 0.30 | 0.43 | 1.22 | 1.00 | 0.99 | 7.89 |
| Total | | 0.48 | 2.40 | 1.50 | 1.24 | 0.26 | 0.31 | 1.90 | 0.05 | 0.31 | 0.52 | 1.55 | 1.00 | 0.99 | 12.51 |

Abbreviations: International Organization of Migration (IOM); Vaccination Program for the US-bound Refugees (VPR); Diphtheria, tetanus, pertussis vaccine (DTP); Tetanus, diphtheria, and acellular pertussis vaccine (Tdap); Tetanus diphtheria vaccine (Td); Hepatitis B adult vaccine (Hep B); Hepatitis B pediatric (HepB Ped); Pentavalent vaccine including diphtheria, tetanus, pertussis, *Haemophilus influenzae* type b (Hib), and hepatitis B antigens (Penta); Bivalent oral polio vaccine (bOPV); Inactivated polio vaccine (IPV); Measles, mumps, rubella vaccine (MMR); Rotavirus vaccine (Rota); Meningococcal conjugate vaccine (MenACWY); Pneumococcal conjugate vaccine (PCV); Varicella vaccine (VAR); Hepatitis A vaccine (HepA); Human papillomavirus vaccine (HPV); Not available (N/A); Middle East and North Africa (MENA); Türkiye and Middle East (TUMA)

Table A3: The number of doses for full vaccination by region for non-IOM sites

**'No VPR' scenario**

| Vaccination location | Region | DTP | Tdap /Td | HepB | HepB Ped | Penta | IPV/ OPV | MMR | Rota | MenACWY | PCV | VAR | HepA | HPV | Total |
|---|---|---|---|---|---|---|---|---|---|---|---|---|---|---|---|
| U.S. | Africa | 0.40 | 2.53 | 1.40 | 1.39 | 0.21 | 0.39 | 1.96 | 0.01 | 0.40 | 0.42 | 1.59 | 1.06 | 1.05 | 12.81 |
| | Asia | 0.35 | 2.50 | 1.77 | 0.98 | 0.25 | 0.22 | 1.94 | 0.03 | 0.25 | 0.51 | 1.47 | 0.82 | 0.84 | 11.93 |
| | MENA/TUME | 0.36 | 2.59 | 1.74 | 1.10 | 0.15 | 0.30 | 1.91 | 0.03 | 0.30 | 0.31 | 1.49 | 0.84 | 0.82 | 11.93 |
| | Europe | 0.24 | 2.66 | 1.96 | 0.89 | 0.15 | 0.13 | 1.83 | 0.05 | 0.29 | 0.30 | 1.42 | 0.70 | 0.67 | 11.28 |
| | Americas | 0.26 | 2.71 | 1.67 | 1.22 | 0.11 | 0.27 | 1.91 | 0.03 | 0.38 | 0.22 | 1.51 | 0.89 | 0.85 | 12.03 |
| | Total | 0.30 | 2.64 | 1.68 | 1.18 | 0.15 | 0.27 | 1.92 | 0.03 | 0.35 | 0.29 | 1.51 | 0.88 | 0.86 | 12.06 |

**'VPR' scenario**

| Vaccination location | Region | DTP | Tdap /Td | HepB | HepB Ped | Penta | IPV/ OPV | MMR | Rota | MenACWY | PCV | VAR | HepA | HPV | Total |
|---|---|---|---|---|---|---|---|---|---|---|---|---|---|---|---|
| Overseas | Africa | 0.05 | 0.97 | 0.54 | 0.61 | 0.12 | 0.15 | 1.12 | 0.01 | N/A | 0.09 | N/A | N/A | N/A | 3.64 |
| | Asia | 0.00 | 0.57 | 0.41 | 0.29 | 0.08 | 0.00 | 0.68 | 0.01 | N/A | 0.08 | 0.36 | N/A | N/A | 2.50 |
| | MENA/TUME | 0.02 | 0.60 | 0.41 | 0.29 | 0.05 | 0.02 | 0.67 | N/A | N/A | N/A | N/A | N/A | N/A | 2.07 |
| | Europe | 0.02 | N/A | 0.34 | 0.12 | 0.05 | 0.03 | 0.56 | N/A | N/A | N/A | N/A | N/A | N/A | 1.12 |
| | Americas | 0.04 | 0.12 | 0.33 | 0.16 | 0.03 | N/A | 0.57 | N/A | N/A | N/A | N/A | N/A | N/A | 1.23 |
| | Total | 0.03 | 0.33 | 0.37 | 0.24 | 0.05 | 0.02 | 0.65 | 0.00 | N/A | 0.02 | 0.05 | N/A | N/A | 1.76 |
| U.S. | Africa | 0.35 | 1.57 | 0.87 | 0.78 | 0.09 | 0.24 | 0.84 | 0.01 | 0.40 | 0.33 | 1.59 | 1.06 | 1.05 | 9.17 |
| | Asia | 0.35 | 1.92 | 1.36 | 0.69 | 0.17 | 0.22 | 1.26 | 0.02 | 0.25 | 0.42 | 1.11 | 0.82 | 0.84 | 9.44 |
| | MENA/TUME | 0.34 | 1.99 | 1.34 | 0.81 | 0.10 | 0.28 | 1.24 | 0.03 | 0.30 | 0.31 | 1.49 | 0.84 | 0.82 | 9.87 |
| | Europe | 0.22 | 2.66 | 1.61 | 0.77 | 0.10 | 0.10 | 1.27 | 0.05 | 0.29 | 0.30 | 1.42 | 0.70 | 0.67 | 10.16 |
| | Americas | 0.22 | 2.59 | 1.34 | 1.07 | 0.08 | 0.27 | 1.34 | 0.03 | 0.38 | 0.22 | 1.51 | 0.89 | 0.85 | 10.79 |
| | Total | 0.27 | 2.31 | 1.31 | 0.94 | 0.10 | 0.26 | 1.27 | 0.03 | 0.35 | 0.27 | 1.46 | 0.88 | 0.86 | 10.29 |
| Total | | 0.30 | 2.64 | 1.68 | 1.18 | 0.15 | 0.27 | 1.92 | 0.03 | 0.35 | 0.29 | 1.51 | 0.88 | 0.86 | 12.06 |

Abbreviations: International Organization of Migration (IOM); Vaccination Program for the US-bound Refugees (VPR); Diphtheria, tetanus, pertussis vaccine (DTP); Tetanus, diphtheria, and acellular pertussis vaccine (Tdap); Tetanus diphtheria vaccine (Td); Hepatitis B adult vaccine (Hep B); Hepatitis B pediatric (HepB Ped); Pentavalent vaccine including diphtheria, tetanus, pertussis, *Haemophilus influenzae* type b (Hib), and hepatitis B antigens (Penta); Bivalent oral polio vaccine (bOPV); Inactivated polio vaccine (IPV); Measles, mumps, rubella vaccine (MMR); Rotavirus vaccine (Rota); Meningococcal conjugate vaccine (MenACWY); Pneumococcal conjugate vaccine (PCV); Varicella vaccine (VAR); Hepatitis A vaccine (HepA); Human papillomavirus vaccine (HPV); Not available (N/A); Middle East and North Africa (MENA);Türkiye and Middle East (TUMA)

### 4. Domestic vaccination costs

In the United States, the vaccine costs for most refugee children under 19 years old are covered by the Vaccines for Children (VFC) program and Medicaid. For adult refugees, most vaccination costs are covered by RMA or Medicaid, as they are eligible for these medical support programs for up to 12 months after their arrival in the United States, provided their date of eligibility for benefits is on or after October 1, 2021 [6].

#### (1) Vaccine costs

##### Child refugees (<19 years old)

The base case domestic vaccine costs for refugee children under 19 years old were estimated using CDC vaccine purchasing costs for the VFC program as of July 1, 2023 (Table A4) [7]. When multiple manufacturers were available for a vaccine, we used the simple average of their costs. The cost of Tdap/Td was calculated as the weighted average of one dose of Tdap and two doses of Td. A 5% vaccine wastage rate was included in the cost estimation [8].

To conduct one-way sensitivity analyses, we considered lower- and upper-bound cost estimates. The lower-bound estimate was identical to the base case, while the upper-bound estimate was based on the private sector purchasing price for each vaccine listed on the VFC websites.

Table A4. Estimated vaccine costs per dose for refugees who are younger than 19 years old in the United States (2023 US dollars)

| Vaccine | Base case | Parameters for one-way sensitivity analyses | |
|---|---|---|---|
| | | Lower-bound | Upper-bound |
| DTaP | $21.97 | $21.97 | $33.48 |
| Tdap/Td | $27.22 | $27.22 | $40.93 |
| HepB Ped | $16.18 | $16.18 | $27.80 |
| Penta | $71.66 | $71.66 | $71.66 |
| IPV | $16.78 | $16.78 | $42.68 |
| MMR | $26.20 | $26.20 | $92.00 |
| Rota | $92.38 | $92.38 | $111.86 |
| MenACWY | $112.05 | $112.05 | $159.91 |

| | | | |
|---|---|---|---|
| PCV | $135.19 | $135.19 | $195.60 |
| VAR | $139.04 | $139.04 | $167.20 |
| HepA | $24.31 | $24.31 | $38.24 |
| HPV | $235.86 | $235.86 | $281.42 |

Abbreviations: Diphtheria, tetanus, and pertussis vaccine (DTaP); Tetanus, diphtheria, and acellular pertussis vaccine (Tdap); Tetanus diphtheria vaccine (Td); Hepatitis B adult vaccine (Hep B); Hepatitis B pediatric (HepB Ped); Pentavalent vaccine including diphtheria, tetanus, pertussis, *Haemophilus influenzae* type b (Hib), and hepatitis B antigens (Penta); Bivalent oral polio vaccine (bOPV); Inactivated polio vaccine (IPV); Measles, mumps, rubella vaccine (MMR); Rotavirus vaccine (Rota); Meningococcal conjugate vaccine (MenACWY); Pneumococcal conjugate vaccine (PCV); Varicella vaccine (VAR); Hepatitis A vaccine (HepA); Human papillomavirus vaccine (HPV); Not available (N/A); Middle East and North Africa (MENA); Türkiye and Middle East (TUMA)

**Adult refugees (≥19 years old)**

The base case costs of adult vaccines were calculated using data from the 2021 MarketScan Medicaid Multi-State Database, focusing on Medicaid beneficiaries aged 19-64 years who had fee-for-service plans. We used the following Current Procedural Terminology (CPT) product codes or drug product name to estimate costs by vaccine [9] (Table A5).

Table A5: Current Procedural Terminology (CPT) product codes and drug product name by vaccine for adult US-bound refugees

| Vaccine | CPT codes | Drug product name |
|---|---|---|
| Tdap | 90715 | Adacel®, Boostrix® |
| Td | 90714 | TDVAX™, Tenivac |
| HepB | 90746 | Heplisav-B™, Engerix-B®, Recombivax HB® |
| MMR | 90707 | M-M-R® II, Priorix |
| VAR | 90716 | Varivax® |
| HPV | 90649, 90650, 90651 | Gardasil®, Gardasil® 9 |

Abbreviations: Tetanus, diphtheria, and acellular pertussis vaccine (Tdap); Tetanus diphtheria vaccine (Td); Hepatitis B adult vaccine (Hep B); Measles, mumps, rubella vaccine (MMR); Varicella vaccine (VAR); Human papillomavirus vaccine (HPV)

We utilized payment data, encompassing Medicaid reimbursement rates and Medicaid beneficiaries' out-of-pocket payments, to estimate the median costs of each vaccine. Records with zero payment records or total payment exceeding $1,000 per vaccine were excluded from the analysis. The median costs from 2021 were adjusted to 2023 levels using the average changes in CDC vaccine purchase prices between 2021 and 2023, as reported on the VFC website [7,10]. Additionally, the cost of Tdap/Td was calculated as the weighted average of one dose of Tdap and two doses of Td.

Estimated vaccine costs for adult refugees are presented in Table A6. The methodology for determining the lower- and upper-bound cost estimates mirrored that used for children's vaccines, including applying the same vaccine wastage rate. However, the base case vaccine cost estimates were not adjusted for vaccine wastage, as the MarketScan payment data reflect the cost of vaccines administered, which inherently accounts for wastage.

Table A6. Vaccine costs per dose for adult refugees (≥19 years old) in the United States (2023 US dollars)

| Vaccine | Base case | Parameters for one-way sensitivity analyses | |
|---|---|---|---|
| | | Lower-bound | Upper-bound |
| Tdap/Td | $37.09 | $22.43 | $37.61 |
| HepB | $75.60 | $50.26 | $95.06 |
| MMR | $91.22 | $64.26 | $93.18 |
| VAR | $162.28 | $111.68 | $167.2 |
| HPV | $273.12 | $187.25 | $281.42 |

Abbreviations: Tetanus, diphtheria, and acellular pertussis vaccine (Tdap); Tetanus diphtheria vaccine (Td); Hepatitis B adult vaccine (Hep B); Measles, mumps, rubella vaccine (MMR); Varicella vaccine (VAR); Human papillomavirus vaccine (HPV)

#### (2) Vaccine administration fee

#### Unit costs of vaccine administration

We used CPT codes 90460 and 90461 to estimate the unit costs of vaccine administration for refugees under 19 years old, and CPT codes 90471 and 90472 to estimate unit costs for adult refugees. For the base case estimates, we calculated the Medicaid vaccine administration fee, following the methodology outlined in Joo et al. (2017) [2]. Since state-specific Medicaid vaccine administration fees were not available, we began by multiplying the 2023 Medicare vaccine administration fee for each state by the proportion of refugee arrivals in each state in FY 2024 [11,12]. We then multiplied these state-specific numbers by the state-specific Medicaid-to-Medicare fee index for primary care in 2019 [13], and summed these estimates across states, as demonstrated in the equation provided by Joo et al. (2017) [2]. For the base case and lower-bound estimates, we assumed zero payment for CPT code 90461, which was not reimbursable for vaccines supplied by VFC in 2023 [14]. The upper-bound estimate was the weighted average of Medicare vaccine administration fees with weights based on state-specific proportions of total US-bound refugee arrivals. All estimates are reported in 2023 US dollars.

Table A7. Estimated vaccine administration fees for refugees in the United States (2023 US dollars)

| | Base case | Parameters for one-way sensitivity analyses | |
|---|---|---|---|
| | | Lower-bound | Upper-bound |
| <19 years old | | | |
| CPT 90460 | $14.78 | $14.78 | $22.60 |
| CPT 90461 | $0 | $0 | $10.16* |
| ≥19 years old | | | |
| CPT 90471 | $13.24 | $13.24 | $20.22 |
| CPT 90472 | $9.49 | $9.49 | $14.51 |

Abbreviation: Current Procedural Terminology (CPT)

* For vaccines given to individuals, less than 19 years of age, the unit cost is calculated depending on the number of components in each vaccine.

**Allowable charge units per dose for administration**

In the United States, vaccine administration fees per dose for infant or children vary based on the number of components in each vaccine. We applied the methodology from Joo et al. (2018) [2] to estimate allowable charge units per dose for administering each vaccine to refugees under 19 years old. For adults, vaccine administration fees depend on the numbers of different vaccines, but not the number of components of individual vaccines, received during a visit and the total number of visits required for completing vaccination. CPT code 90471 is used for the administration of the first single or combination vaccine for individuals aged 19 and older, while CPT code 90472 is used for the administration of each additional vaccine and must be billed in conjunction with CPT code 90471 during a single visit. We assumed that three visits would be necessary to complete the vaccination series for US-bound adult refugees. Therefore, three instances of CPT code 90471 were required, with all remaining vaccine doses associated with CPT code 90472.

Total vaccine administration fees for an individual under 19 years old were estimated by multiplying the number of units, unit costs (Table A7), and the number of doses required for full vaccination (Table A2 and A3), separately for CPT 90460 and 90461. For adults, total vaccine administration fees were calculated by adding costs associated with CPT 90471 multiplied by three to account for three visits (Table A7) to the unit costs associated with CPT 90472 for the total number of doses minus three (Table A7). Additional details are provided in Joo et al. (2018) [2].

### 5. Overseas vaccination costs

We used FY 2024 IOM budget data to estimate the overseas vaccination costs associated with VPR. These costs included both vaccine costs and program costs. The projected number of overseas vaccine doses delivered through VPR were previously reported in Tables A2 and A3. For vaccine-specific costs, we estimated the cost per dose for each vaccine by region and IOM/non-IOM site based on budget data from over 80 countries where VPR operated in FY2024 (Table A8). The estimated costs include a 5% vaccine wastage rate and a 7% overhead cost.

Program costs consisted of two types of variable costs a) those that varied based on the total number of doses administered through VPR and b) those that varied by the number of refugees. We also included fixed costs, which remained constant regardless of the number of doses or refugees. Variable costs associated with the number of refugees included expenses for Hepatitis B surface antigen (HBsAg) screening (Table A9). Variable costs related to total number of doses administered and included administration fees charged by non-IOM sites and all other non-vaccine operational expenses, excluding HBsAg screening costs, pregnancy test costs, and antiparasitic drug costs from the IOM budget data. We did not include pregnancy test costs or antiparasitic drug costs in this analysis because those activities would be executed independently of VPR. Fixed costs encompassed expenses such as office operations, staff salaries, and cold chain maintenance (Table A10). Each program cost item included a 7% of overhead charge. These calculations use the full medical caseload of 177,400. To estimate the cost per 100,000 refugee cohort, we assumed the same distribution of refugees arriving by region and IOM versus non-IOM sites.

Table A8: Overseas vaccine costs per dose with VPR (2023 US dollars)

| | Region | DTP | Tdap/ Td | HepB | HepB Ped | Penta | IPV/ bOPV | MMR | Rota | MenACWY | PCV | VAR |
|---|---|---|---|---|---|---|---|---|---|---|---|---|
| IOM | Africa | $0.20 | $0.15 | $1.41 | $0.27 | $2.81 | $0.14 | $3.11 | $21.31 | $32.10 | $3.10 | $28.77 |
| | Asia | $4.43 | $2.22 | $6.63 | $5.95 | $31.67 | $9.74 | $14.75 | $33.23 | N/A | $22.68 | $23.20 |
| | MENA/TUME | $4.22 | $0.10 | $3.85 | $2.95 | $8.21 | $1.35 | $9.56 | N/A | N/A | $33.17 | N/A |
| | Europe | $45.88 | N/A | N/A | N/A | $44.42 | $38.71 | $21.97 | N/A | N/A | N/A | N/A |
| | Grand Total | $2.69 | $0.62 | $3.46 | $2.07 | $12.34 | $1.73 | $7.65 | $24.66 | $32.10 | $4.32 | $27.52 |
| Non-IOM | Africa | $12.95 | $7.67 | $8.63 | $1.65 | $12.88 | $2.43 | $8.60 | $21.23 | N/A | $3.10 | N/A |
| | Asia | $17.37 | $9.92 | $6.64 | $4.91 | $20.25 | $10.55 | $10.75 | $14.98 | N/A | $33.17 | $18.25 |
| | MENA/TUME | $7.13 | $5.74 | $12.25 | $8.14 | $30.60 | $6.44 | $12.08 | N/A | N/A | N/A | N/A |
| | Europe | $28.53 | N/A | $45.29 | $29.52 | $33.64 | $40.36 | $29.71 | N/A | N/A | N/A | N/A |
| | America | $40.21 | $17.18 | $19.60 | $16.15 | $72.96 | N/A | $33.03 | N/A | N/A | N/A | N/A |
| | Grand Total | $32.91 | $9.74 | $15.40 | $9.40 | $38.03 | $4.77 | $22.53 | $17.03 | N/A | $21.08 | $18.25 |

Abbreviations: International Organization of Migration (IOM); Vaccination Program for the US-bound Refugees (VPR); Diphtheria, tetanus, pertussis vaccine (DTP); Tetanus, diphtheria, and acellular pertussis vaccine (Tdap); Tetanus diphtheria vaccine (Td); Hepatitis B adult vaccine (Hep B); Hepatitis B pediatric (HepB Ped); Pentavalent vaccine including diphtheria, tetanus, pertussis, *Haemophilus influenzae* type b (Hib), and hepatitis B antigens (Penta); Bivalent oral polio vaccine (bOPV); Inactivated polio vaccine (IPV); Measles, mumps, rubella vaccine (MMR); Rotavirus vaccine (Rota); Meningococcal conjugate vaccine (MenACWY); Pneumococcal conjugate vaccine (PCV); Varicella vaccine (VAR); Hepatitis A vaccine (HepA); Human papillomavirus vaccine (HPV); Not available (N/A); Middle East and North Africa (MENA); Türkiye and Middle East (TUMA)

Table A9: Estimated variable program costs from IOM's FY 2024 VPR budget

| | Region | Medical caseload (A) | Doses (B) | HBsAg screening (C) | Operational costs (D) | HBsAg screening per person, (E)=(C)/(A) | Operational costs per dose, (F)=(D)/(B) |
|---|---|---|---|---|---|---|---|
| IOM | Africa | 34,000 | 178,679 | $60,027 | $51,947 | $1.77 | $0.34 |
| | Asia | 14,700 | 71,063 | $107,515 | $30,082 | $7.31 | $0.88 |
| | MENA/TUME | 21,000 | 84,218 | $42,834 | $20,132 | $2.04 | $0.22 |
| | Europe | 3,000 | 1,701 | $21,400 | $2,415 | $7.13 | $23.99 |
| | Grand Total | 72,700 | 335,660 | $231,776 | $104,576 | $3.19 | $0.54 |
| Non-IOM | Africa | 9,900 | 36,064 | $52,965 | $7,918 | $5.35 | $0.22 |
| | Asia | 15,300 | 38,180 | $255,570 | $27,611 | $16.70 | $0.72 |
| | MENA/TUME | 15,000 | 30,992 | $80,357 | $69,978 | $5.36 | $2.26 |
| | Europe | 2,500 | 2,809 | $32,100 | $23,726 | $12.84 | $8.45 |
| | America | 62,000 | 76,508 | $1,256,116 | $278,200 | $20.26 | $3.64 |
| | Grand Total | 104,700 | 184,553 | $1,677,107 | $407,433 | $16.02 | $2.21 |

Abbreviations: Fiscal year (FY); Vaccination Program for the US-bound Refugees (VPR); International Organization of Migration (IOM); Hepatitis B surface Antigen (HBsAg); Middle East and North Africa (MENA); Türkiye and Middle East (TUMA)

Table A10: Estimated fixed program costs from IOM's FY 2024 VPR budget

| | Region | Medical caseload, (A) | Office cost, (B) | Staff cost, (C) | Cold chain maintenance cost, (D) | Fixed cost per person, $(E) = \frac{(B)+(C)+(D)}{(A)}$ |
|---|---|---|---|---|---|---|
| IOM | Africa | 34,000 | $51,947 | $363,509 | $10,791 | $12.54 |
| | Asia | 14,700 | $30,082 | $314,095 | $20,899 | $24.84 |
| | MENA/TUME | 21,000 | $20,132 | $360,087 | $3,766 | $18.29 |
| | Europe | 3,000 | $2,415 | $150,936 | N/A | $51.12 |
| | Grand Total | 72,700 | $104,576 | $1,188,627 | $35,457 | $18.28 |
| Non-IOM | Africa | 9,900 | $2,460 | $19,682 | N/A | $2.24 |
| | Asia | 15,300 | $16,141 | $74,354 | $13,121 | $6.77 |
| | MENA/TUME | 15,000 | $3,728 | $29,821 | N/A | $2.24 |
| | Europe | 2,500 | $621 | $4,970 | N/A | $2.24 |
| | America | 62,000 | $57,779 | $217,269 | $16,050 | $4.70 |
| | Grand Total | 104,700 | $80,729 | $346,096 | $29,171 | $4.36 |

Fiscal year (FY); Vaccination Program for the US-bound Refugees (VPR); International Organization of Migration (IOM); Not available (N/A); Middle East and North Africa (MENA); Türkiye and Middle East (TUMA)

### 6. One-way sensitivity analysis with domestic vaccination costs

US domestic vaccination costs consist of vaccine-specific prices as well as non-specific vaccine administration fees. We considered ranges of estimated values to conduct one-way sensitivity analyses. We defined the baseline domestic vaccination cost estimates to represent the most likely estimates for US-bound refugees, who will be covered by RMA or Medicaid for up to 12 months after their arrival [6]. The upper-bound estimate was obtained by assuming that refugees are covered by private insurance and pay more for vaccinations through their private insurance. The lower-bound estimate uses the lowest possible costs. For instance, CDC vaccine purchasing costs for the VFC program for adult vaccines are slightly lower than the estimates from Medicaid claims data, so we used VFC program costs for adult vaccines in place of estimates derived from Medicaid claims data.

The results from the one-way sensitivity analysis using ranges of domestic vaccination costs are shown in Table A11. The range of net cost savings per person varies from $115.3 to $245.9 (base case: $147.0). The range of net cost savings per 100,000 refugee cohort in 2024 varies from $11.5 million to $24.6 million (base case: $14.7 million).

Table A11: Results from one-way sensitivity analysis with range of domestic vaccination price by region (2023 US dollars)

| | Region | Net cost savings per person, 2023 US dollar | | | Total net cost savings per 100,000 refugee cohort in 2024, million 2023 US dollar | | |
|---|---|---|---|---|---|---|---|
| | | Lower-bound | Baseline | Upper-bound | Lower-bound | Baseline | Upper-bound |
| IOM | Africa | $290.0 | $332.5 | $524.7 | $5.6 | $6.4 | $10.1 |
| | Asia | $195.4 | $250.5 | $401.7 | $1.6 | $2.1 | $3.3 |
| | MENA/TUME | $159.8 | $208.8 | $336.7 | $1.9 | $2.5 | $4.0 |
| | Europe | -$51.9 | -$44.1 | -$20.4 | -$0.09 | -$0.1 | -$0.03 |
| | Americas | N/A | N/A | N/A | N/A | N/A | $0.00 |
| | Total | $219.1 | $264.7 | $422.8 | $9.0 | $10.8 | $17.3 |
| Non-IOM | Africa | $147.2 | $182.8 | $307.8 | $0.82 | $1.0 | $1.7 |
| | Asia | $118.3 | $145.4 | $224.1 | $1.02 | $1.3 | $1.9 |
| | MENA/TUME | $68.7 | $95.3 | $155.3 | $0.58 | $0.8 | $1.3 |
| | Europe | $1.4 | $20.0 | $56.1 | $0.00 | $0.03 | $0.08 |

| | | | | | | | |
|---|---|---|---|---|---|---|---|
| | Americas | $3.5 | $21.3 | $63.3 | $0.12 | $0.75 | $2.2 |
| | Total | $43.2 | $65.3 | $122.8 | $2.6 | $3.9 | $7.3 |
| Grand Total | | $115.3 | $147.0 | $245.9 | $11.5 | $14.7 | $24.6 |

Abbreviations: International Organization of Migration (IOM); Middle East and North Africa (MENA); the Republic of Türkiye and Middle East (TUME); Not available (N/A)

Notes: Lower-bound estimates used the CDC Vaccines for Children (CDC VFC) purchasing price for adults. Lower-bound estimates for child vaccines are as same as the baseline estimates (CDC VFC children's purchasing price). Upper-bound estimates used the private sector price from the CDC VFC website for both adult and child vaccines. The '-' signs represent net costs as opposed to net cost savings for a region.

### 7. Sub-model analyses

We conducted three sub-model analyses to address uncertainties related to VPR implementation overseas. We selected key factors that could influence costs under the 'VPR' scenario to develop sub-model analyses. The analyses include: (1) Sub-model 1: historical vaccine records documentation–incorporates additional net cost savings resulting from the documentation of historical vaccine records overseas, (2) Sub-model 2: alternative Americas vaccine cost–estimation based on potentially lower vaccine prices for the Americas , and (3) Sub-model 3: bOPV utilization and IPV re-vaccination accounts for pre-departure bOPV use in some countries, which would increase domestic costs for IPV re-vaccination in the United States.

#### (1) Sub-model 1: historical vaccine records documentation–incorporates additional cost savings resulting from the documentation of historical vaccine records overseas

Through VPR, overseas panel sites also request that US-bound refugees present any historical vaccination records at the time of health assessment. Panel sites then document valid historical vaccines, which would count towards completing the VPR schedule. Each refugee's full immunization record, including valid historical and panel-provided doses, is transmitted to the United States as part of the electronic medical record. These records are accessible to domestic refugee health providers in the United States, preventing unnecessary revaccination after resettlement.

VPR recorded an average of 0.99 historical doses per person in 2023 (e.g., vaccine doses recorded on an individual refugee's historical vaccination records which were not provided by the panel site) Of the total, 13% were COVID-19 vaccines and 69% were vaccines considered in the analysis. The remaining 18% of reported historical doses include vaccines that are not present on the US ACIP schedule, such as yellow fever, and therefore would not count towards completing the US schedule. The average number of historical doses per person for the vaccines considered in our analysis was 0.68 (Table A12). Our overseas budget estimate already accounted for the reduced number of vaccine doses provided by panel sites when accounting for valid, documented vaccine histories (e.g., an estimated 0.7 doses may be included in the budget analysis under the assumption that about 3 in 10 refugees would have documentation of previous vaccination); however, our baseline analysis did not account for the potential cost savings associated with overseas historical vaccine documentation (i.e., we did not consider potential savings in the United States based on the assumption that the number of doses needed would be reduced by the refugees' documented vaccine histories). Based on the reported distribution of documented historical doses, we estimated potential post-arrival net cost savings of $57.4 per dose in the US by assuming that the refugees would not be revaccinated. The net cost savings per person would be $39.3, which would result in an additional $4.3 million in net cost savings relative to our baseline estimates for 100,000 refugee cohort. The total net cost savings associated with VPR after accounting for historical doses would be $19.0 million per 100,000 refugee cohort.

Table A12: Additional cost savings due to the records of historical vaccine doses from VPR, (2023 US dollars)

| | Region | Average cost per US dose, (A) | Documented historical doses per person, (B) | Average savings due to documented historical doses per person, (C)=(A)×(B) | Total net cost savings, accounting for documented historical doses per person, (D) | Medical caseload for 100,000 refugee cohort, (E) | Additional cost savings due to documented historical doses for 100,000 refugee cohort (million US dollars), (F)=(C)×(E) | Total net cost savings with documented historical doses for 100,000 refugee cohort (million US dollars)*, (G)=(D)×(E) |
|---|---|---|---|---|---|---|---|---|
| IOM | Africa | $56.6 | 0.64 | $36.0 | $368.6 | 19,166 | $0.7 | $7.1 |
| | Asia | $57.7 | 0.59 | $34.3 | $284.8 | 8,286 | $0.3 | $2.4 |
| | MENA/TUME | $57.5 | 0.35 | $19.9 | $228.7 | 11,838 | $0.2 | $2.7 |
| | Europe | $58.1 | 0.64 | $37.0 | -$7.1† | 1,691 | $0.1 | -$0.01† |
| | Americas | N/A | N/A | N/A | N/A | N/A | N/A | N/A |
| | Total | $57.1 | 0.57 | $32.8 | $297.5 | 40,981 | $1.3 | $12.1 |
| Non-IOM | Africa | $56.9 | 0.57 | $32.7 | $215.4 | 5,581 | $0.2 | $1.2 |
| | Asia | $57.9 | 0.58 | $33.7 | $179.1 | 8,625 | $0.3 | $1.5 |
| | MENA/TUME | $57.8 | 0.98 | $56.8 | $152.2 | 8,455 | $0.5 | $1.3 |
| | Europe | $58.5 | 1.08 | $63.1 | $83.1 | 1,409 | $0.1 | $0.1 |
| | Americas | $57.6 | 0.97 | $55.8 | $77.2 | 34,949 | $3.5 | $2.7 |
| | Total | $57.6 | 0.95 | $55.0 | $120.3 | 59,019 | $2.0 | $6.8 |
| Grand Total | | $57.4 | 0.68 | $39.3 | $186.3 | 100,000 | $4.3 | $19.0 |

Abbreviations: Vaccination Program for the US-bound Refugees (VPR); International Organization of Migration (IOM); Middle East and North Africa (MENA); the Republic of Türkiye and Middle East (TUME); Not available (N/A)

Note: *Column G indicates the cost savings associated with VPR.

†The '-' signs represent net cost.

**(2) Sub-model 2: alternative Americas vaccine cost–estimation based on potentially lower future vaccine prices for the Americas**

We considered regional variation in vaccine costs in the main analysis. As a sub-model, we considered the potential for IOM to access Pan American Health Organization (PAHO) Revolving Fund vaccine prices for calendar year 2024 [15] (Table A13) to reduce costs in the Americas region. Note that this PAHO price is not currently an option for VPR. At present, IOM, via VPR, provides vaccines for the Americas through non-IOM sites, resulting in relatively higher vaccine costs in this region. If IOM were able to access lower prices through PAHO and distribute those vaccines to non-IOM sites, VPR costs may decline. When comparing the alternative cost estimates using PAHO Revolving Fund vaccine prices to the FY2024 IOM budget costs for the Americas, the alternative estimates are significantly lower (Table A13).

Table A13: Estimated vaccine costs for Americas using PAHO Revolving Fund's vaccine prices (US dollar)

| | | Presentation (doses per vial) | PAHO posted price (per dose) | Estimated cost* (per dose) | FY2024 IOM budget cost |
|---|---|---|---|---|---|
| DTP | DTP | 10 | $0.19 | $0.26 | $40.21 |
| | DTaP | 1 | $19.29 | $25.72 | |
| | Average | N/A | $9.74 | $12.99 | |
| Td | Td (reduced antigen) | 10 | $0.13 | $0.17 | $17.18 |
| HepB | Hepatitis B Adult | 1 | $0.87 | $1.16 | $19.60 |
| | Hepatitis B Adult | 10 | $0.33 | $0.45 | |
| | Average | N/A | $0.60 | $0.80 | |
| HepB Ped | Hepatitis B Pediatric | 1 | $0.60 | $0.81 | $16.15 |
| Penta | DTP-HepB-Hib (Liquid) | 1 | $1.19 | $1.59 | $72.96 |
| MMR | MMR (Jeryl-Lynn strain of Mumps) | 1 | $5.30 | $7.07 | $33.03 |
| | MMR (Zagreb strain of Mumps) | 1 | $3.56 | $4.75 | |
| | | 5 | $1.78 | $2.37 | |
| | | 10 | $1.71 | $2.28 | |
| | Average | N/A | $3.09 | $4.12 | |

Abbreviations: Pan American Health Organization (PAHO); Fiscal year (FY); International Organization of Migration (IOM); Diphtheria, tetanus, pertussis vaccine (DTP); Tetanus diphtheria vaccine (Td); Hepatitis B adult vaccine (Hep B); Hepatitis B pediatric (HepB Ped); Pentavalent vaccine including diphtheria, tetanus, pertussis, *Haemophilus influenzae* type b (Hib), and hepatitis B antigens (Penta); Measles, mumps, rubella vaccine (MMR); Not available (N/A)
Note: *Estimated prices were multiplied first by 5% to account for wastage, and then by an additional 27% to account for shipping, and transporting vaccines (including doses lost to wastage) from government sites to panel sites (20%) and overhead (7%).

As a result, we would anticipate further VPR-associated cost savings for refugees arriving from the Americas. The FY2024 VPR-associated estimated net cost savings, with a caseload of 34,949 from the Americas region for a 100,000 refugee cohort, are \$0.75 million using the baseline assumptions, but could increase to \$1.8 million when using PAHO Revolving Fund vaccine prices (Table A14).

Table A14: Estimated costs of full vaccination per person, except for influenza and COVID-19 vaccines, with and without the VPR **in Americas** (2023 US dollars)

| | 'No VPR' scenario, cost per person (A) | 'VPR' scenario, cost per person | | | Net cost savings per person, (E)=(A)-(B) | Total number of doses per person | Number of overseas (VPR) doses per person | Net cost savings per overseas dose, (E)/(G) | Total net cost savings (N=34,949 refugees per 100,000 refugee cohort) |
|---|---|---|---|---|---|---|---|---|---|
| | | Total, (B)=(C)+(D) | Overseas, (C) | United States, (D) | | | | | |
| Estimates with FY2024 IOM budget cost | $1,067.9 | $1,046.5 | $62.7 | $983.9 | $21.3 | 11.9 | 1.2 | $17.3 | $0.75 million |
| Estimates using PAHO Revolving Fund's vaccine prices | $1,067.9 | $1,016.6 | $32.7 | $983.9 | $51.3 | | | $41.6 | $1.8 million |

Abbreviations: Fiscal year (FY); Vaccination Program for the US-bound Refugees (VPR); International Organization of Migration (IOM); Pan American Health Organization (PAHO)

Notes: With current VPR prices, overseas vaccination costs in the Americas are $2.2 million, resulting in $0.75 million in net cost savings compared to the 'No VPR' scenario. Using PAHO pricing, overseas vaccination costs in the Americas would be $1.1 million, resulting in $1.8 million in cost savings compared to the 'No VPR' scenario. Total net cost savings were based on a medical caseload of 34,949 refugees from the Americas region for a 100,000 refugee cohort.

(3) **Sub-model 3: bOPV utilization and IPV re-vaccination to account for use of bOPV pre-departure use in some countries, which requires domestic costs for IPV re-vaccination after arrival in the United States**

In the main analysis, we assumed that US-bound refugees who received any type of polio vaccine overseas would not require revaccination with IPV in the United States, irrespective of whether they received bOPV (bivalent, oral polio vaccine) or IPV (trivalent, inactivated polio vaccine). However, some VPR sites only have access to bOPV. Thus, as of 2024, VPR currently provides bOPV at certain sites in Africa, MENA/TUME, and Asia. In accordance with ACIP recommendations, individuals who received bOPV overseas would be revaccinated with IPV after arrival in the United States. Thus, the estimated net cost savings from the main analysis may be overestimated. To address this issue, we examined the worst-case scenario in which all individuals who received polio vaccines overseas would be re-vaccinated with IPV in the US. Although the worst-case scenario is unlikely to happen, the estimates from sub-model 3 provide the maximum range of additional costs associated with IPV revaccination in the United States.

Our base case domestic IPV vaccination cost per dose is $31.56, which includes $16.78 for IPV vaccine and $14.78 for the vaccine administration fee. According to the FY 2024 IOM budget, 12,421 doses of polio vaccine would be administered to US-bound refugee children prior to departure. After pro-rating to a cohort of 100,000 refugees, this would correspond to 7,002 doses. If all these doses were re-administered after their resettlement in the United States, the estimated cost would be $0.22 million, leading to a decrease in net cost savings associated with VPR relative to our baseline estimate.

Even under this worst-case scenario, the decrease in net cost savings is minimal representing around 1.5% of the total net cost savings associated with VPR. Furthermore, the administration of polio vaccines overseas provides the additional benefit of preventing polio outbreaks and mitigating the risk of polio case importations by US-bound refugees. The impact of VPR on polio outbreak prevention will be further discussed in the subsequent section.

### 8. Impact of VPR on vaccine preventable disease outbreaks

VPR reduces the risk of vaccine-preventable disease (VPD) outbreaks among US-bound refugees, depending on which vaccines are delivered and the risk of VPD outbreaks in each country. Given the concerning current global measles and polio outbreak situations, we will focus on measles and polio outbreaks to estimate the potential benefits of vaccinating US-bound refugees prior to departure in the event an outbreak occurs. Because measles and polio pose significant public health risks, outbreaks among US-bound refugees could lead to substantial travel delays if cases are identified.

While VPR would reduce risks associated with other vaccine-preventable disease outbreaks, including meningococcal disease, diphtheria, pertussis, varicella, and influenza among US-bound refugees, these vaccines are not always available in countries where outbreaks occur (e.g., MenACWY and varicella vaccines) or were not included in this analysis (e.g., influenza vaccines). There are also limited data on the cost of VPD outbreaks within the US, especially the costs associated with refugees who arrived infected or potentially exposed to these diseases.

We estimate that VPR has the potential to prevent four to five large-scale travel delays resulting from measles cases among US-bound refugees who are processed in countries with ongoing outbreaks. In addition, we considered the costs associated with one to two large-scale travel delays and post-arrival monitoring related to polio outbreaks in overseas camps or localities directly hosting US-bound refugees each year under the 'No VPR' scenario. These delays typically occur to allow implementation of pre-travel outbreak measures, including monitoring and vaccination, for US-bound refugees. In the absence of VPR, all vaccines would have to be procured and administered at the pre-travel stage, at higher cost and logistic effort than would be required during routine vaccination. We also would expect that despite these reactive outbreak control measures there would still be an increased risk of measles or polio importation into the United States, necessitating domestic response activities. The estimated annual total costs of responding to measles and polio outbreaks in the "No VPR" scenario would amount to $1.1million, with a range of $0.5 million to $10.3 million (Table A15). Therefore, VPR could potentially save an estimated $1.1 million annually by reducing costs associated with measles and polio outbreaks responses both overseas and in the United States.

Table A15: Summary of estimated annual total outbreak response costs associated with vaccine preventable disease outbreaks among US-bound refugees (2023 US dollars)

| | Base case | Lower-bound | Upper-bound |
|---|---|---|---|
| Measles, (A) | $446,013 | $140,228 | $9,120,424 |
| Polio, (B) | $700,826 | $350,413 | $1,168,043 |
| Total, (C)=(A)+(B) | $ 1,146,839 | $490,641 | $10,228,468 |

**Measles outbreaks**

Measles outbreaks are among the most common VPD outbreaks in countries hosting US-bound refugees, with cases reported in approximately 60 such countries as of FY2024. Of these, 50% are in sub-Saharan Africa, 22% are in Europe, 18% are in the MENA region, and the remaining 10% are in Asia. Before implementation of VPR, measles outbreaks in host countries frequently caused significant travel delays for US-bound refugees. For example, in 2004, outbreaks of measles, rubella, and varicella in camps hosting Liberian refugees in Côte d'Ivoire required the delivery of MMR vaccinations to around 3,000 US-bound refugees and resulted in travel delays of over six months for 5,400 to 8,000 refugees, as well as a death in a child waiting to resettle to the US and the birth of an infant severely disabled by congenital rubella syndrome [16].

Based on the FY2024 global VPD outbreak landscape, we assumed that there would be approximately four measles outbreaks per year occurring within overseas camps or localities directly hosting US-bound refugees (Table A16). The lower- and upper- bound estimates were calculated by adding and subtracting 50% of baseline. Under the ‘no VPR’ scenario, all these situations would result in travel delays while overseas outbreak measures are implemented. This would include time for vaccines to be procured and administered before travel to the United States, as well as a quarantine period before travel. We estimated the base case overseas response costs per outbreak based on Coleman et al. (2017)—which included costs for rescheduling US-bound refugees for later flights, vaccinating resettling refugees, and treating a hospitalized case ($88,751 in 2023 US dollars, accounting for 1,500 US-bound refugees affected in that report) (Table A16) [17]. We used this figure as the baseline estimate per outbreak. The lower- and upper-bound estimates were calculated by adding and subtracting 25% of baseline costs. The estimated overseas response costs associated with measles outbreaks were $355,005 per year (range: $133,127-$931,889).

Table A16: Estimated annual overseas response costs associated with measles outbreaks (2023 US dollars)

| | Baseline | Lower-bound | Upper-bound |
|---|---|---|---|
| Number of overseas refugee camps hosting US-bound refugees with measles outbreak per year, (A) | 4 | 2 | 6 |
| Overseas response costs per measles outbreak, (B) | $88,751 | $66,564 | $155,315 |
| Overseas measles outbreak response costs per year, (C) = (A) × (B) | $355,005 | $133,127 | $931,889 |

Among these two to six  measles outbreaks per year, we would expect at least one imported measles case per year to occur in an unvaccinated US-bound refugee, leading to a US domestic measles outbreak investigation with its associated costs. The cost of US outbreak investigation would be additive to the costs of overseas outbreak measures described previously. The lower-bound estimate of the imported measles cases per year was 0.2, based on data from 2000 to 2010, during which two reported measles importations occurred among US-bound refugees [18]. We assumed that the upper-bound estimate was two imported measles cases per year. This estimate may increase considering the increasing frequency and spread of global measles outbreaks.

The baseline estimate was based on Coleman et al. (2017), which calculated the costs of a multi-state outbreak response initiated by an index case of a Burmese refugee from Malaysia, resulting in seven domestic measles cases [17]. The baseline estimate per measles importation was $91,007 in 2023 US dollars (Table A3), including costs associated with labor from the CDC and state health departments, measles testing, vaccination, outpatient treatment and hospitalization.

The lower-bound estimated domestic response costs per measles importation were estimated using Coleman et al. (2012), which assessed the costs associated with a single measles-infected US-bound refugee in 2010 [18]. The estimate cost was $35,507 in 2023 US dollars, covering costs related to labor, medical care, transportation, and vaccination (Table A3) [18]. This importation resulted in 44 contacts but no additional cases in Kentucky. However, as of 2024, the risk of measles outbreaks spreading in the US may be higher than it was in the Kentucky example, due to pockets of unvaccinated US residents [19]. The upper-bound estimate was based on Pike et al. (2021), a recently published study assessing the societal costs of measles response activities for an outbreak in Clark County, Washington that resulted in 72 cases between December 31, 2018, and April 26, 2019 [20]. The upper-bound estimate per measles importation was $4,094,268 in 2023 US dollars, including costs associated with the public health response to the outbreak, productivity loss for measles patients and for exposed quarantined persons who lacked presumptive evidence of immunity as well as for caregivers, and direct medical costs. The estimate is greater than the upper-bound estimate ($1,063,936) from Pike et al. (2020), which reviewed 10 published manuscripts between 2001 and 2018 estimating the costs associated with measles outbreak responses [21].

Under the "No VPR" scenario, the estimated annual domestic response costs associated with measles importations by US-bound refugees were $91,007 in 2023 US dollars, with a range of $7,101 to $8,188,535 (Table A17).

Table A17: Estimated annual domestic response costs associated with measles importations by US-bound refugees (2023 US dollars)

| | Base case | Lower-bound | Upper-bound |
|---|---|---|---|
| Number of imported measles cases by US-bound refugees per year, (A) | 1 | 0.2 | 2 |
| Domestic response costs per measles importation, (B) | $91,007 | $35,507 | $4,094,268 |
| Domestic measles importation response costs per year, (C) = (A) × (B) | $91,007 | $7,101 | $8,188,535 |

The estimated total annual response costs associated with measles outbreaks, including both overseas and domestic response costs, were $446,013 in 2023 US dollars (range: $140,228 - $9,120,424) (Table A18).

Table A18: Estimated annual total response costs associated with measles outbreaks (2023 US dollars)

| | Base case | Lower-bound | Upper-bound |
|---|---|---|---|
| Overseas, (A) | $355,005 | $133,127 | $931,889 |
| Domestic, (B) | $91,007 | $7,101 | $8,188,535 |
| Total, (C)=(A)+(B) | $446,013 | $140,228 | $9,120,424 |

**Polio outbreaks**

Polio outbreaks have been reported in 35 countries hosting US-bound refugees in FY2024, of which 29 are in sub-Saharan Africa. There have been no imported polio cases among US-bound refugees to-date as of FY2024. However, under the 'No VPR' scenario, we expected that there would be approximately one or two polio outbreaks per year in refugee camps or other localities directly hosting US-bound refugees (Table A19), which would lead to large-scale travel delays of more than 4 weeks while overseas outbreak measures are implemented. Additionally, the outbreaks could trigger domestic response activities including active monitoring of refugees who arrived before the polio outbreak was identified.

There is a single published manuscript estimating the costs of responding to a polio outbreak in a camp hosting US-bound refugees [22]. The outbreak occurred between October and December 2006 in Dadaab, northeastern Kenya [22]. After arriving in the United States, 944 US-bound refugees from these camps received IPV vaccines and were placed under active polio surveillance by state and local health departments, including 34 state health departments [22]. Among these refugees, 163, who were in Nairobi awaiting departure, were screened for polio symptoms and administered one dose of trivalent oral polio vaccine (tOPV) before traveling to the US [22]. Note that, as of 2024, most global polio outbreaks are caused by cVDPV2, which is no longer covered by OPV. As a result, a more expensive and less readily available polio vaccine (i.e., IPV) is required for polio outbreak response [23]. The travel of 1,200 US-bound refugees in Dadaab was delayed by several months while they received two doses of monovalent oral poliovirus vaccine type 1 (mOPV1) [22]. The total cost of this response was $309,283 in 2006 US dollars [22]. These costs included overseas response costs, such as labor, transportation (e.g., nonrefundable flight tickets), vaccination, and stool testing for polio [22]. The US domestic response costs included labor, vaccination, stool testing, interpreter services, and other costs [22].

We adjusted the estimate to 2023 US dollars using the Consumer Price Index for all urban consumers (CPI-U) from the U.S. Bureau of Labor Statistics [24], resulting in a base case estimate of $467,217 per polio outbreak in 2023 U.S. dollars (Table A19). The estimated total annual response costs related to polio outbreaks among US-bound refugees were $700,826, with a range of $350,413 to $1,168,043.

Table A19: Estimated total annual response costs associated with polio outbreaks (2023 US dollars)

| | Base case | Lower-bound | Upper-bound |
|---|---|---|---|
| Number of overseas refugee camps hosting US-bound refugees with polio outbreak per year, (A) | 1.5 | 1 | 2 |
| Overseas and domestic response costs per polio outbreak, (B) | $467,217 | $350,413 | $584,022 |
| Polio outbreak response costs per year, (C) = (A)×(B) | $700,826 | $350,413 | $1,168,043 |

### 9. Costs comparison by age group

Table A20: Costs of full vaccination per person, except for influenza and COVID-19 vaccines, with and without the VPR, **by age** (2023 US dollars).

| | Region | 'No VPR' scenario, (A) | 'VPR' scenario | | | Net cost savings, (E)=(A)-(B) | Percent cost reduction, (E)/(A) | Total number of doses | Number of overseas doses | Net cost saving per overseas dose, (E)/(G) |
|---|---|---|---|---|---|---|---|---|---|---|
| | | | Total, (B)=(C)+(D) | Overseas, (C) | United States, (D) | | | | | |
| <19 years old | IOM | $1,263.06 | $1,013.40 | $64.45 | $948.95 | $249.66 | 20% | 15.1 | 5.2 | $48.06 |
| | Non-IOM | $1,190.13 | $1,152.43 | $56.86 | $1,095.57 | $37.70 | 3% | 14.8 | 1.9 | $20.27 |
| | Total | $1,222.25 | $1,089.96 | $58.96 | $1,031.00 | $132.29 | 11% | 14.9 | 3.3 | $39.75 |
| ≥19 years old | IOM | $1,014.26 | $733.26 | $39.88 | $693.38 | $281.00 | 28% | 10.0 | 4.0 | $69.58 |
| | Non-IOM | $994.34 | $906.78 | $52.51 | $854.27 | $87.55 | 9% | 9.9 | 1.7 | $51.93 |
| | Total | $1,391.15 | $841.38 | $48.76 | $792.62 | $160.59 | 12% | 10.0 | 2.6 | $62.07 |

Abbreviations: Vaccination Program for US-bound Refugees (VPR); International Organization for Migration (IOM); Not Available (N/A)

Table A21: Costs of full vaccination per person, except for influenza and COVID-19 vaccines, excluding the Americas (2023 US dollars)

| IOM status | 'No VPR' scenario, (A) | 'VPR' scenario | | | Net cost savings, (E)=(A)-(B) | Percent cost reduction, (E)/(A) | Total number of doses | Number of overseas doses, (F) | Net cost saving per overseas dose, (E)/(F) |
|---|---|---|---|---|---|---|---|---|---|
| | | Total, (B)=(C)+(D) | Overseas, (C) | Domestic, (D) | | | | | |
| IOM | $1,136.9 | $872.2 | $52.2 | $820.0 | $264.7 | 23% | 12.5 | 4.6 | $57.3 |
| non-IOM | $1,095.2 | $966.0 | $42.5 | $923.6 | $129.1 | 12% | 11.9 | 2.5 | $51.0 |
| Total | $1,121.4 | $906.9 | $48.6 | $858.4 | $214.5 | 19% | 12.4 | 3.8 | $55.8 |
| Joo et al. (2018) | $785.2 | $559.3 | $46.2 | $513.1 | $225.9 | 29% | 13.1 | 6.4 | $35.4 |

Notes: The figures in the row from Joo et al. (2018) are sourced from Table 1 of that study and reported in 2015 US dollars [2]. The difference in the total number of doses between 2024 and 2017 may be due to changes in age distribution and the introduction of the pentavalent vaccine, which combines three vaccines into one dose in 2023. In 2018, we assumed all refugees received two doses of available vaccines overseas. In 2024, we used FY2024 budget data, which is more realistic than the hypothetical data used in 2018. The US vaccination costs without VPR increased by around 40%. The overseas vaccination costs increased by around 5%.

3. US Centers for Disease Control and Prevention. 2024 recommended immunization for birth through 6 years old 2024.

4. US Centers for Disease Control and Prevention. 2024 Recommended Immunizations for adults aged 19 years and older, 2024.

5. US Centers for Disease Control and Prevention. 2024 recommended immunization for children 7-18 years old 2024.

6. Administration for Children & Families, US Department of Health and Human Services. HHS Extends Refugee Assistance in Historic Eligibility Expansion, 2022.

7. US Centers for Disease Control and Prevention. VFC CDC Vaccine Price List as of July 1, 2023, 2023.

8. Zhou F, Shefer A, Wenger J, et al. Economic evaluation of the routine childhood immunization program in the United States, 2009. *Pediatrics* 2014;133:577-585.

9. Immunization Information Systems Support Branch-Informatics, Centers for Disease Control and Prevention. CPT codes mapped to CVX codes, 2024.

10. US Centers for Disease Control and Prevention. VFC CDC Vaccine Price List as of July 1, 2021, 2021.

11. Centers for Medicare & Medicaid Services. Physician fee schedule, 2023.

12. Refugee Admissions, Bureau of Population, Refugees, and Migration, US Department of State. Refugee Arrivals by State and Nationality, 2024.

13. KFF. Medicaid-to-Medicare Fee Index: Timeframe 2019.

14. American Academy of Pediatrics. FAQs on the Medicaid payment increase for primary care and immunization administration services & updates to the vaccines for children program regional maximum charges, 2012.

15. Pan America Health Organization. Vaccine Prices 2024, 2024.

16. Plotinsky RN, Talbot EA, Kellenberg JE, et al. Congenital rubella syndrome in a child born to Liberian refugees: clinical and public health perspectives. *Clin Pediatr (Phila)* 2007;46:349-355.

17. Coleman MS, Burke HM, Welstead BL, et al. Cost Analysis of Measles in Refugees Arriving at Los Angeles International Airport from Malaysia. *Hum Vaccin Immunother* 2017;13:1084-1090.

18. Coleman MS, Garbat-Welch L, Burke H, et al. Direct costs of a single case of refugee-imported measles in Kentucky. *Vaccine* 2012;30:317-321.

19. US Centers for Disease Control and Prevention. Vaccination Coverage and Exemptions among Kindergartners, 2024.

20. Pike J, Melnick A, Gastanaduy PA, et al. Societal Costs of a Measles Outbreak. *Pediatrics* 2021;147.

21. Pike J, Leidner AJ, Gastanaduy PA. A Review of Measles Outbreak Cost Estimates From the United States in the Postelimination Era (2004-2017): Estimates by Perspective and Cost Type. *Clin Infect Dis* 2020;71:1568-1576.

22. Centers for Disease Control and Prevention. U.S.-incurred costs of wild poliovirus infections in a camp with U.S.-bound refugees--Kenya, 2006. *MMWR Morb Mortal Wkly Rep* 2008;57:232-235.

23. World Health Organization. Statement of the fortieth meeting of the Polio IHR Emergency Committee, 2024.

24. Bureau of Labor Statistics, United States Department of Labor. Consumer Price Index for all urban consumers.
